# Development of PROWalk: a functional mobility person-reported outcome measure based on the PROMIS^®^ adult physical function item bank

**DOI:** 10.1101/2025.08.21.25334089

**Authors:** Sutton B. Richmond, Laura Jacobs, Caleb Kumar, Yvonne Rumsey, Carole A. Tucker, Lars Oddsson

## Abstract

**Introduction:** Assessing an individual’s physical capacity to move independently and safely to perform activities of daily living (i.e., functional mobility) can be approached through various modalities, including performance-based physiological tests, observational assessments, and self-report questionnaires. The purpose of this study was to establish a patient-reported outcome measure (PROM), termed PROWalk^TM^, based on the PROMIS Item Bank 2.0 – Physical Function, to assess perceived mobility challenges, specifically related to gait and balance function, and evaluate its performance in comparison to the Patient-Specific Functional Scale (PSFS) within a population of individuals with sensory peripheral neuropathy.

**Methods:** The PROWalk PROM was developed with expert clinician input as a custom short form composed of selected items from the Item Bank. Participants with lower extremity peripheral neuropathy who had been prescribed a wearable sensory prosthesis were administered the PROWalk and the PSFS, a perceived functional mobility assessment via telecommunications at baseline and six weeks after wearing the sensory prosthesis.

**Results:** Both measures displayed significant increases (i.e., improvements) over time with large effect sizes and upward linear trends for each instrument between baseline and six weeks. When normalized, the measures showed no bias, large limits of agreement, and no normalized change between the instruments.

**Discussion:** The PROWalk PROM detected significant improvements in perceived functional mobility over time and showed strong temporal stability and suitability for standardized group-level comparisons, which were similar to the PSFS. Overall, the PROWalk appears to be a valid PROM for capturing improvements in self-reported functional mobility related to gait and balance function.

## 1. Introduction

Functional mobility refers to an individual’s physical capacity to move independently and safely in order to perform activities of daily living (e.g., transferring, housekeeping, shopping, transportation) (1). The assessment of functional mobility can be approached through various modalities, including performance-based physiological tests, observational assessments, and patient-reported outcome measures (PROM) (2). Specifically, PROM provides patient-centered insights into individual functional ability, capturing real-world participation, perceived capability, and symptom impact, which performance measures conducted in standardized settings may miss (3). The PROM construct is supported by robust regulatory guidance (4), validated measurement frameworks such as the Patient-Reported Outcomes Measurement Information System (PROMIS^®^) (5, 6), and implementation standards enabling precise, scalable, and even remote assessment (7). If PROMs are used alongside performance tests, they offer complementary information that enhances ecological validity and ensures the evaluation reflects outcomes meaningful to patients (3).

One widely used instrument is the Patient-Specific Functional Scale (PSFS), which enables individuals to identify up to five activities they find challenging due to a health condition. Each activity is self-rated based on the individual’s perceived level of difficulty. The PSFS has demonstrated strong psychometric properties, including excellent test–retest reliability (ICC = 0.848) and good construct validity, evidenced by moderate correlations with established functional outcome measures (8, 9). Finally, the PSFS is highly responsive to clinical change and offers individualized insights into patient-prioritized outcomes, making it especially useful in physical therapy and rehabilitation settings (9-11). However, the PSFS is limited by the absence of standardized metrics, the lack of discrete mobility domain identification, and its vulnerability to variability and biases. To address these limitations, we aimed to develop the PROWalk^TM^, a standardized PROM for assessing gait and balance in adults with sensory peripheral neuropathy (SPN). Ostensibly, an alternative tool that retains the personalized strengths of the PSFS while improving standardization, domain-specific resolution, and clinical applicability.

The Patient-Reported Outcomes Measurement Information System (PROMIS, development funded by the National Institutes of Health, comprises a comprehensive suite of 300 rigorously validated self-report instruments designed to assess a broad spectrum of health domains, including physical, mental, and social well-being (12, 13). Among these, the PROMIS physical function item bank have been shown to be valid and reliable to measure change in research and clinical applications. PROMIS item banks support computer adaptive test (CAT) administration and have defined standard 4- and 8-item fixed-length, or short, forms that provide T-scores. Given that PROMIS item banks are calibrated using the item response theory (IRT), any set of items from a bank can be selected to provide “custom” short forms consisting of user-selected items that are scored on the same metric (14).

The PROMIS physical function bank consists of 165 IRT-calibrated items using a 5-point Likert scale, with most items scored on a difficulty response scale where “5” corresponded to “without any difficulty” and “1” to “unable to perform the activity.” To date, no systematic efforts have been made to establish specific PROMIS physical function items that relate closely to the critical domains of walking and balance function, i.e., functional mobility. The motivation for our work was to develop an 8 – 10-item custom short form of PROMIS physical function items specific to balance and gait-related functional impairments for use in adults with lower extremity conditions/ impairments. To address this gap, we systematically developed a patient-reported outcome instrument, termed PROWalk, from the PROMIS Item Bank v2.0 – Physical Function, specifically designed to assess mobility-related challenges. This study outlines the conceptual and methodological foundation of PROWalk. Further, we report its convergent validity with the PSFS within a population of individuals with SPN who present with gait and balance issues.

## 2. Materials and methods

### 2.1. Participants

A convenience sample of fifty-one participants (height: 178.8 ± 8.3 cm, mass: 96.6 ± 19.9 kg, and age: 76 ± 4 years, 4 female) provided verbal informed consent for this study. All participants were required to provide verbal consent, be age-qualified (≥65 years), and have been prescribed Walkasins^®^, a wearable sensory prosthesis designed to replace part of the lost plantar sensory nerve function and improve mobility in individuals with SPN (15, 16). SPN is a prevalent global health concern characterized by damage to the sensory nerves responsible for detecting touch, pressure, and vibration, which are critical inputs required for maintaining postural stability and balance (17-19). As a common manifestation of polyneuropathy, SPN affects approximately 50% of individuals with diabetes (20) and is frequently observed in cancer patients undergoing treatment with neurotoxic chemotherapeutic agents, resulting in chemotherapy-induced peripheral neuropathy (21, 22). The sensory deficits associated with SPN can lead to significant impairments in mobility, heightened risk of falls, and a decline in functional independence and overall quality of life (23, 24). Participants were excluded if their physical condition had changed significantly since baseline due to an injury, hospitalization, or other adverse event that disrupted their typical daily activities. Advarra IRB served as the Institutional Review Board and approved study procedures and consent in accordance with the Declaration of Helsinki.

### 2.2. Protocol

Data for this investigation were collected as part of a company-led initiative by RxFunction, Inc., under the Outcomes Program, which was developed to gain a deeper understanding of the challenges faced by individuals with SPN. The study design incorporated a baseline touchpoint followed by a post-intervention assessment conducted approximately six weeks later (ranging from −2 to +7 days relative to the target follow-up date).

Participants in this study used Walkasins, a wearable sensory prosthesis designed to partially restore plantar sensory function and improve mobility in individuals with SPN (15, 16). At each assessment timepoint, participants completed a standardized battery of both quantitative and qualitative questionnaires, which evaluated key domains including demographic and anthropometric data, overall health status, and physical functionality, particularly mobility-related outcomes. To ensure procedural consistency, all follow-up assessments were administered in the same sequence as the baseline evaluations and recorded in REDCap, an electronic data capture system (25, 26).

The assessment battery was anchored by two perceived functional mobility measures: the PSFS and the newly developed PROWalk instrument (*development details below*). All examinations were conducted by a trained rater, who followed standardized administration protocols. For the PSFS, participants were asked to identify up to three activities that they were either unable to perform or found difficult to perform due to gait and balance impairments associated with peripheral neuropathy. Participants were explicitly prompted to reflect on how these impairments affected their daily functional tasks. Once activities were identified, the rater documented each activity, and participants were then asked to rate the degree of difficulty associated with performing each one using an 11-point numeric scale, where: “0” was indicative of being “Unable to perform” and “10” indicated the participant was “Able to perform at the same level as prior to the onset of the problem or injury” (12, 13). Individual activity scores were then averaged to produce a composite functional status score, ranging from “0” to “10,” with higher scores indicating better perceived function (8).

The development of the PROWalk followed a structured four-step process to ensure conceptual clarity, clinical relevance, and psychometric rigor: (i) *Item Compilation and Initial Screening*: Five independent reviewers evaluated the 165 items from the PROMIS^®^ Item Bank v2.0 – Physical Function (available at healthmeasures.net), each selecting a minimum of eight items, consistent with the typical length of PROMIS short forms, that were explicitly related to gait and postural control. Item selection for the PROWalk questionnaire was informed by qualitative insights gathered during patient follow-up. Specifically, we incorporated activities frequently mentioned by patients in their responses to the PSFS during Outcomes calls. These patient-reported activities reflected real-world challenges and priorities directly voiced by users of the prosthesis and were instrumental in shaping a questionnaire that is both relevant and representative of the lived experience of the clinical population. (ii) *Cross-Rater Consolidation*: The initial selections were consolidated into a list of 24 candidate items, which were then cross-referenced across raters to identify areas of agreement and divergence. (iii) *Item Reduction*: The item pool was refined to 12 items by excluding those that (a) overlapped conceptually or semantically with other items, (b) were not applicable to individuals with impaired mobility, or (c) lacked relevance to functional activities influenced by mobility limitations. (iv) *Final Item Selection and Conceptual Framework*: A final set of 9 items (see **Table 1**) was agreed upon by all four reviewers, based on the following conceptual criteria:

**Table 1.**
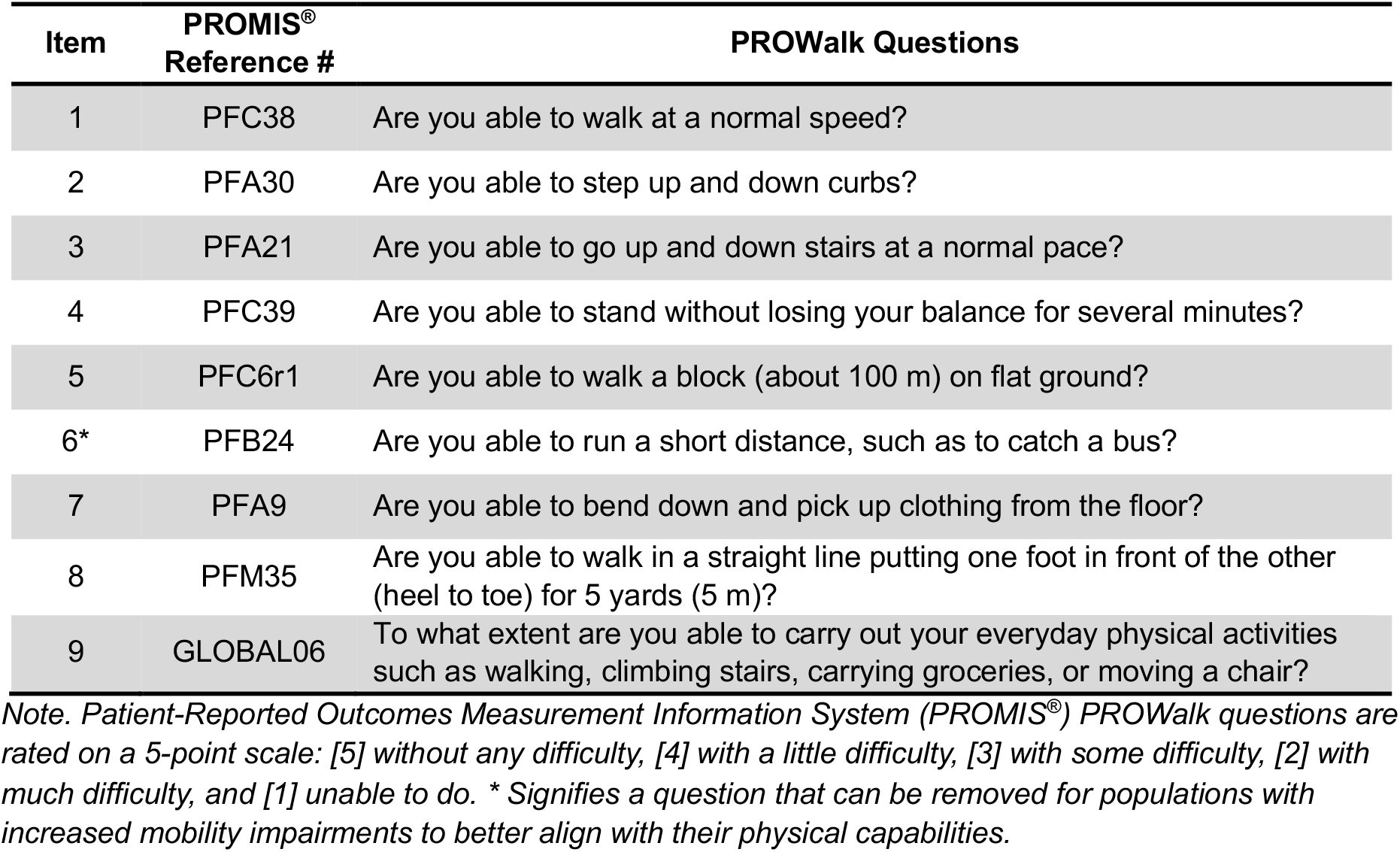
PROWalk Custom Short Form Items from the PROMIS^®^ Physical Function Item Bank.

- Items should capture a broad range of physical functioning to mitigate floor and ceiling effects observed in prior user populations.
- Items must include both standing and walking activities, with a greater emphasis on walking tasks.
- Selected items should reflect activities that participants have identified as meaningful during Outcomes Program interviews, specifically those they aspire to improve.

This process yielded a targeted, patient-relevant instrument tailored to capture core domains of perceived functional mobility, with a focus on gait and balance. An additional single item was included on overall physical functioning (GLOBAL06) was also self-rated on a 5-point Likert scale where “5” corresponded to “completely” and “1” to “not at all” (**Table 1**). For all telehealth communications, raters were trained and utilized a standardized script to enhance the reproducibility of the protocol.

### 2.3. Statistics

Descriptive statistics were computed to summarize the distribution of scores from the PROWalk and PSFS questionnaires. Data normality was assessed using the Shapiro-Wilk test to determine suitability for parametric testing (27). The PROWalk scores at both baseline and follow-up did not significantly (p > 0.05) deviate from normality. However, the PSFS scores at follow-up deviated from normal distribution (W = 0.938, p =.010), supporting the use of non-parametric methods in subsequent analyses. T-scores were generated from the HealthMeasures Scoring Service for PROWalk performances (14). In the system, we selected “Custom Short Form,” “PROMIS,” “Adult,” and “Physical Function.” The item bank from which we had selected the items was PROMIS v2.0 - Physical Function, and we used the default calibration sample (PROMIS 1 Wave 1 with Extension) because the logs showed that all nine items we had chosen were used to calculate the T-scores (28, 29). Z-scores were calculated by subtracting the population mean from each individual raw score (PSFS) or T-score (PROWalk) and dividing the result by the standard deviation, thereby standardizing values for cross-participant comparison. To evaluate changes over time and compare results between instruments, paired t-tests (Wilcoxon signed-rank) were conducted on both the raw data and the standardized (z-score) scores of the PROWalk and PSFS. Effect sizes were interpreted as trivial (<0.20), small (0.20-0.49), medium (0.5-0.79), or large (>.80) (30).

Spearman’s correlation coefficient (*rho*) was calculated to examine the relationship between the two questionnaires and evaluate the strength and direction of linear association. To assess the relationship between changes in mobility-specific patient-reported outcomes and perceived functional status, a correlation analysis was conducted between the z-score standardized change scores from the PROWalk and PSFS instruments. Correlation coefficients were interpreted using commonly accepted thresholds: *rho* = 0.0-0.19 (very weak), 0.20–0.39 (weak), 0.40–0.59 (moderate), 0.60-0.79 (strong), and 0.80-1.0 (very strong) (31). All analyses were performed using JASP (University of Amsterdam, Amsterdam, Netherlands, Version 0.19.3) with risk of type I error set at α=.05. Graphical representations were derived using GraphPad Prism 10 (GraphPad Software, La Jolla, CA, Version 10.5.0), and results are presented with 95% confidence intervals where applicable.

## 3. Results

Descriptive data for each dependent measure are provided in **Table 2**. Participants (N = 51) demonstrated significant improvements on both outcome measures over time (scattered plotted individually in *Supplemental Material Figure 1*), with large effect sizes registered (**Table 2**). Furthermore, both instruments showed a positive correlation over time. **Figure 1** illustrates this relationship, showing an upward linear trend for each instrument (**Figures 1A** and **1B**), with the PROWalk evaluation exhibiting stronger temporal consistency (*rho* = 0.684, *p* < 0.001, **Figure 1A**).

**Table 2.** Descriptives (mean ± standard deviation) and summary of the test of differences between baseline and follow-up.

| Raw Scores | Baseline | Follow-Up | Statistic | z | p | Effect Size |
| --- | --- | --- | --- | --- | --- | --- |
| PROWalk T-scores | 32.85 ± 4.45 | 36.36 ± 4.31 | 77.500 | -5.406 | <0.001 | -0.878 |
| PSFS | 3.10 ± 1.70 | 5.95 ± 2.17 | 4.000 | -6.053 | <0.001 | -0.993 |

*Note. Patient-Specific Functional Scale (PSFS). For the Wilcoxon test, effect sizes are given by the matched rank biserial correlation. Significant mean differences ( $p$ < .05)*
*Note. PROWalk 5-point Likert scale: "5" corresponded to "completely" and "1" to "not at all"*
*Note. PSFS 11-point numeric scale: "0" was indicative of being "Unable to perform" and "10" indicated the participant was "Able to perform at the same level as prior to the onset of the problem or injury"*
*Note: A negative matched rank biserial correlation in the configured paired differences (timepoint 1 – timepoint 2) is indicative of improved physical function between timepoints.*

**Figure 1.**
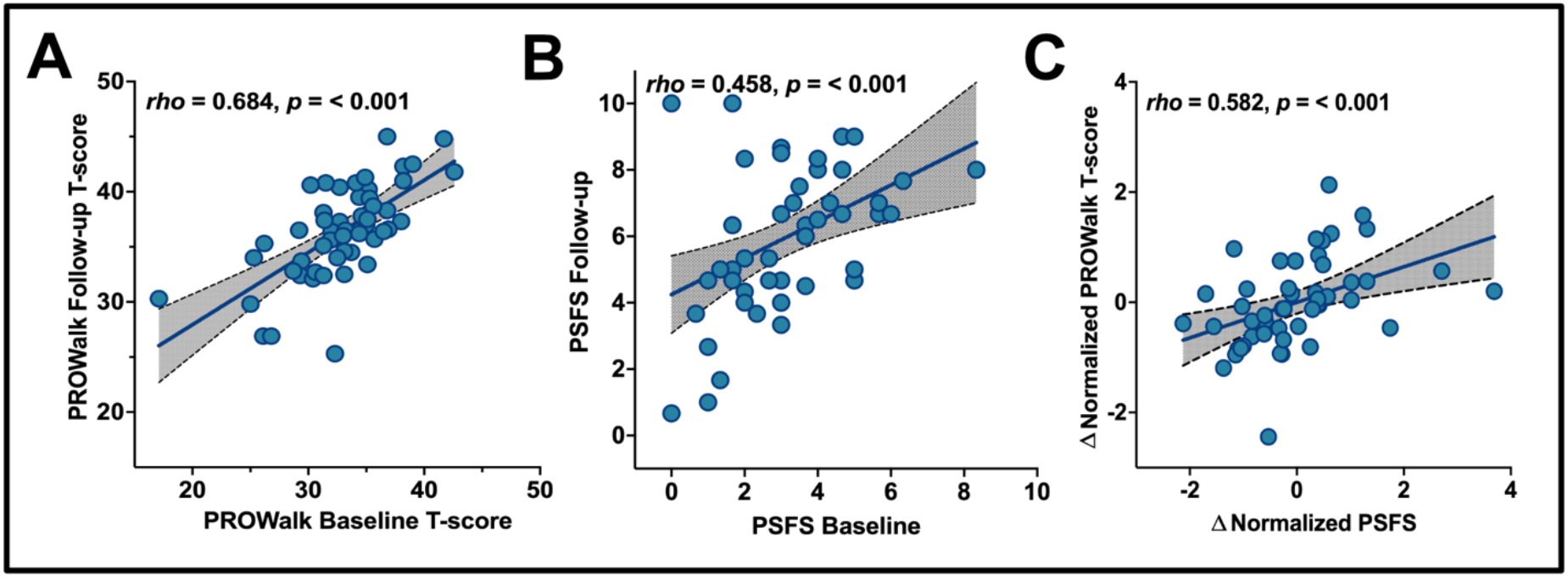
Correlations between trials and the change in scoring between instruments in persons with sensory peripheral neuropathy. Significant, positive correlations were identified for the A.) PROWalk (*rho* = 0.684, *p* = <0.001), B.) PSFS (*rho* = 0.458, *p* = <0.001), and C.) the change (Δ) in normalized instrument scores over time (*rho* = 0.582, *p* = <0.001).

When the instrument data were normalized, the mean difference (bias) between the PROWalk and PSFS Z-scores was approximately zero (M = −3.92 × 10^-11^), indicating no systematic measurement bias between the two instruments at baseline (**Figure 2A**). However, the limits of agreement (LOA) ranged from −2.238 to 2.238. The follow-up agreement analysis revealed similar outcomes (**Figure 2B**). Furthermore, there was no significant difference (*p* = 0.725) in the normalized change between the instruments, with negligible effect sizes (0.057). A moderate positive correlation (*rho* = 0.582, *p* < 0.001) was exhibited between the change scores in PROWalk and PSFS after normalization (**Figure 1C**).

**Figure 2.**
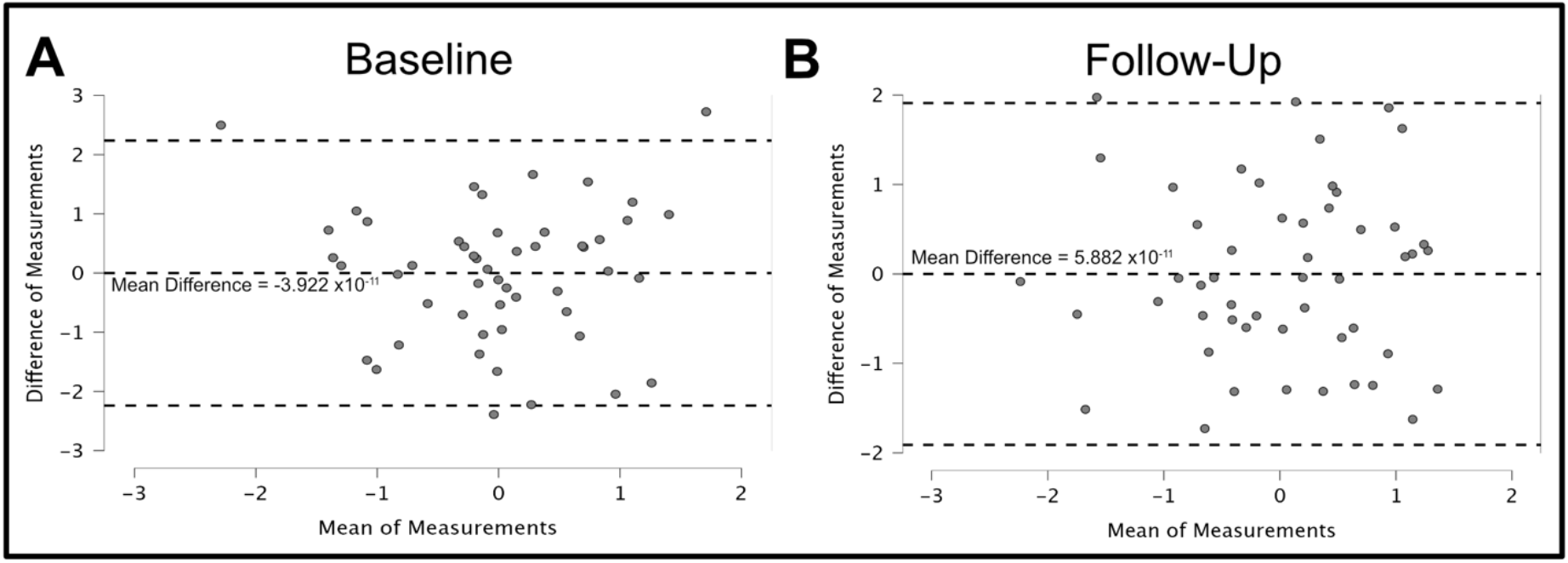
Bland-Altman plot identifying the agreement between the normalized PROWalk and PSFS instruments at (**A**) baseline and (**B**) follow-up timepoints.

## 4. Discussion

This study evaluated the performance of the novel PROWalk instrument alongside the PSFS to assess perceived functional mobility in individuals with SPN who present with gait and balance issues. Individuals demonstrated significant improvements in perceived functional mobility, as measured by both instruments, following six weeks of using the wearable sensory prosthesis. Additionally, scores on the two instruments were moderately to strongly correlated from baseline to follow-up. The absence of significant score changes within each instrument over time, when normalized, further supports the validity of the PROWalk as a robust measure of perceived functional mobility. Although no systematic bias was observed between the instruments at either time point, the wide LOA suggests that individual-level discrepancies may restrict their interchangeability.

Both instruments effectively detected improvements in perceived functional mobility over time. The PSFS is well-established as a valid, reliable, and responsive outcome measure across a range of clinical populations (8, 10, 11). It has also been shown to be appropriate for between-group comparisons and for assessing change at the group level (32). Prior research has demonstrated the PSFS’s utility in tracking progressive improvement with therapeutic intervention (8), a finding corroborated in the present study. Although the PSFS exhibited slightly greater responsiveness, reflected by a larger effect size (−0.993) compared to that of the PROWalk (−0.878), this difference was minimal. The slightly greater sensitivity of the PSFS may reflect its heightened responsiveness to individual-level functional changes compared to the PROWalk. However, further research is warranted to explore this distinction more fully.

Although the PROWalk demonstrated greater temporal stability between timepoints (*rho* = 0.684, *p* < 0.001; **Figure 1A**) compared to the PSFS (*rho* = 0.458, *p* < 0.001; **Figure 1B**), the reliability of each instrument could not be formally assessed due to the interventional nature of the study design. Nevertheless, the stronger temporal consistency observed in the PROWalk suggests that this novel instrument not only captures intervention-induced improvements but may also offer greater stability in tracking generalized perceived functional mobility over time. When normalized change scores were compared across instruments, a significant positive correlation emerged, indicating that participants who reported greater improvement on the PROWalk also tended to report greater improvement on the PSFS. Despite some individual-level variability, the overall pattern reflects a consistent direction of perceived functional improvement across both measures. While descriptive results indicated an upward trend in both instrument scores from baseline to follow-up, the statistical tests did not detect significant within-subject changes when scores were standardized (Δ*Z*). This outcome suggests that while participants may report perceived improvement in function and quality of life, the magnitude of this change, once normalized, is statistically indistinct across the two instruments. The results also suggest the two instruments may reflect changes in related but non-identical constructs of functional status and perceived health.

While no systematic bias was observed between instruments in either interpretation of the baseline or follow-up Bland-Altman plots, the degree of individual-level disagreement underscores a limitation in their interchangeability for assessing perceived functional mobility. These findings are consistent with previous work by Abbott and colleagues on the PSFS (32), supporting the notion that such instruments may be appropriate for group-level analyses, but should be interpreted with caution when applied to individual-level clinical decision-making. The broader LOA shown in both Bland-Altman plots further emphasize the need for context-specific instrument selection. Specifically, the PSFS may offer greater sensitivity in detecting individualized perceived functional impairments, whereas the PROWalk may provide a more standardized assessment of general perceived functional status.

This study has several important limitations. First, the study design did not include a control group (e.g., individuals without Walkasins use or neurotypical participants), limiting our ability to validate the findings and precluding formal reliability analysis between instruments. Additionally, the participant sex distribution was skewed toward males, and future evaluations should better evaluate the female effects. The presence of a potentially performance-enhancing intervention further hindered the ability to assess test–retest reliability across timepoints. Additionally, the absence of a clinically validated physical performance measure, such as the Functional Gait Assessment (33, 34) or another established “gold standard” assessment of functional mobility, limited our capacity to benchmark the tested instruments. While the PSFS is recognized as a valid, reliable, and responsive outcome measure (8), its highly individualized nature may constrain its utility for direct comparison with the PROWalk. Nonetheless, the selection of the PSFS was considered the most appropriate choice given the opportunity to use it in a telehealth-based setting. Future research should focus on validating the PROWalk against more responsive and quantitative physical performance (e.g., gait) measures to better establish its reliability and clinical utility in assessing perceived functional mobility. Moreover, studies should aim to validate the PROWalk in diverse clinical populations, such as individuals post-stroke or those living with Parkinson’s disease, to broaden its applicability and generalizability.

Lastly, the PROWalk is designed to assess the full spectrum of mobility; however, more impaired individuals may find question six, which was originally included to minimize any ceiling effects (**Table 1**), unrelatable, therefore potentially limiting the aggregate score relevance across samples. Consequently, we recommend limiting the assessment to the eight applicable questions (i.e., PROWalk-8) in populations where mobility is more severely impacted. In the supplemental material (*Supplemental Table 1 and Figure 2*), we have provided an identical analysis to the full PROWalk, displaying similar outcomes in the PROWalk-8.

In conclusion, the PROWalk demonstrated a comparable ability to detect significant improvements in perceived functional mobility over time, along with strong temporal stability and suitability for standardized group-level comparisons, similar to the PSFS. Correlational analyses further supported moderate concordance in change detection, underscoring the complementary utility of the two instruments. Overall, the PROWalk emerges as an effective and promising measure for capturing perceived improvements in functional mobility. Continued investigation into its clinical and research applications is warranted to establish its validity and utility further.

## Data Availability

All data produced in the present work are contained in the manuscript

## Acknowledgements

Development of the REDCap system, used in this research, was supported by the National Institutes of Health’s National Center for Advancing Translational Sciences, grant UM1TR004405. The content is solely the responsibility of the authors and does not necessarily represent the official views of the National Institutes of Health’s National Center for Advancing Translational Sciences.

