## Supplemental Material for "Development of PROWalk: a functional mobility person-reported outcome measure based on the PROMIS^®^ adult physical function item bank"

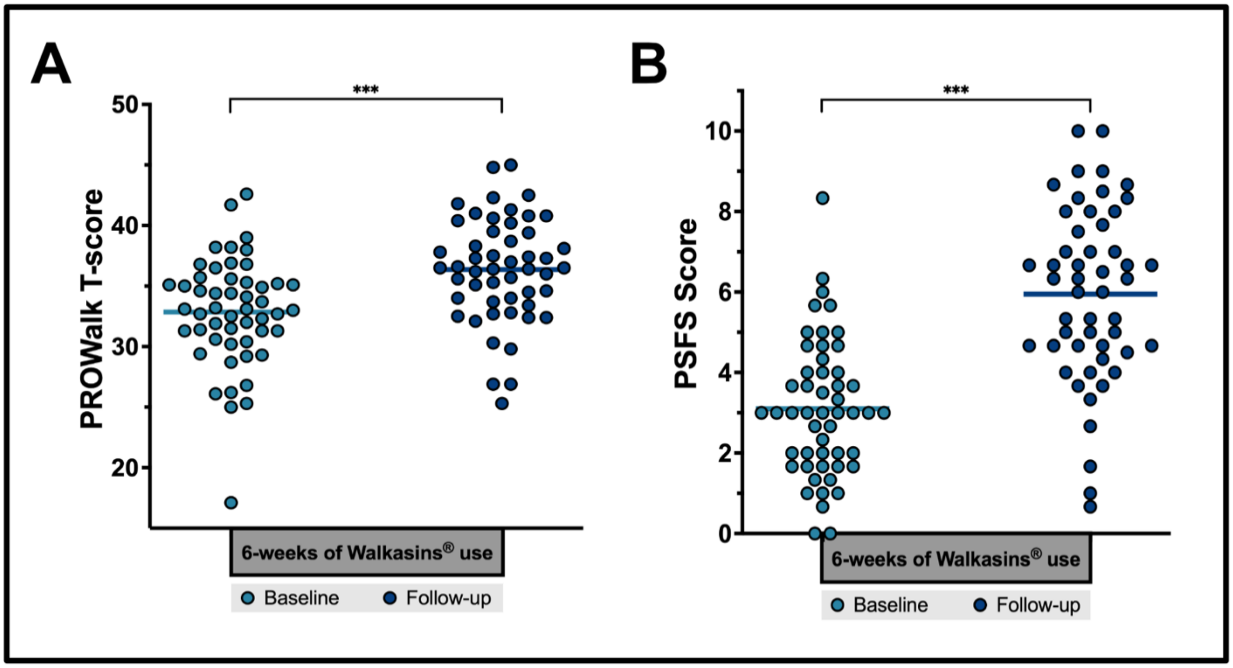


**Supplemental Figure 1. A)** PROWalk performance before (32.85 ± 4.45) and six weeks after (36.36 ± 4.31) wearing Walkasins® in persons with sensory peripheral neuropathy. **B)** Patient-Specific Functional Scale (PSFS) performance before (3.10 ± 1.70) and six weeks after (5.95 ± 2.17) wearing Walkasins® in the same sample. The triple star indicates significance at the 0.1% level.

**Supplemental Table 1. Descriptives (mean ± standard deviation) and summary of the test of differences between baseline and follow-up of the PROWalk-8**

| Measure | Baseline | Follow-Up | Statistic | z | *p* | *Effect Size* |
|---|---|---|---|---|---|---|
| PROWalk-8 T-scores | 32.91 ± 4.48 | 36.75 ± 4.72 | 71.500 | -5.464 | <0.001 | -0.888 |

*Note. For the Wilcoxon test, effect sizes are given by the matched rank biserial correlation. Significant mean differences (p < .05).*
*Note: A negative matched rank biserial correlation in the configured paired differences (timepoint 1 – timepoint 2) is indicative of improved physical function between timepoints.*

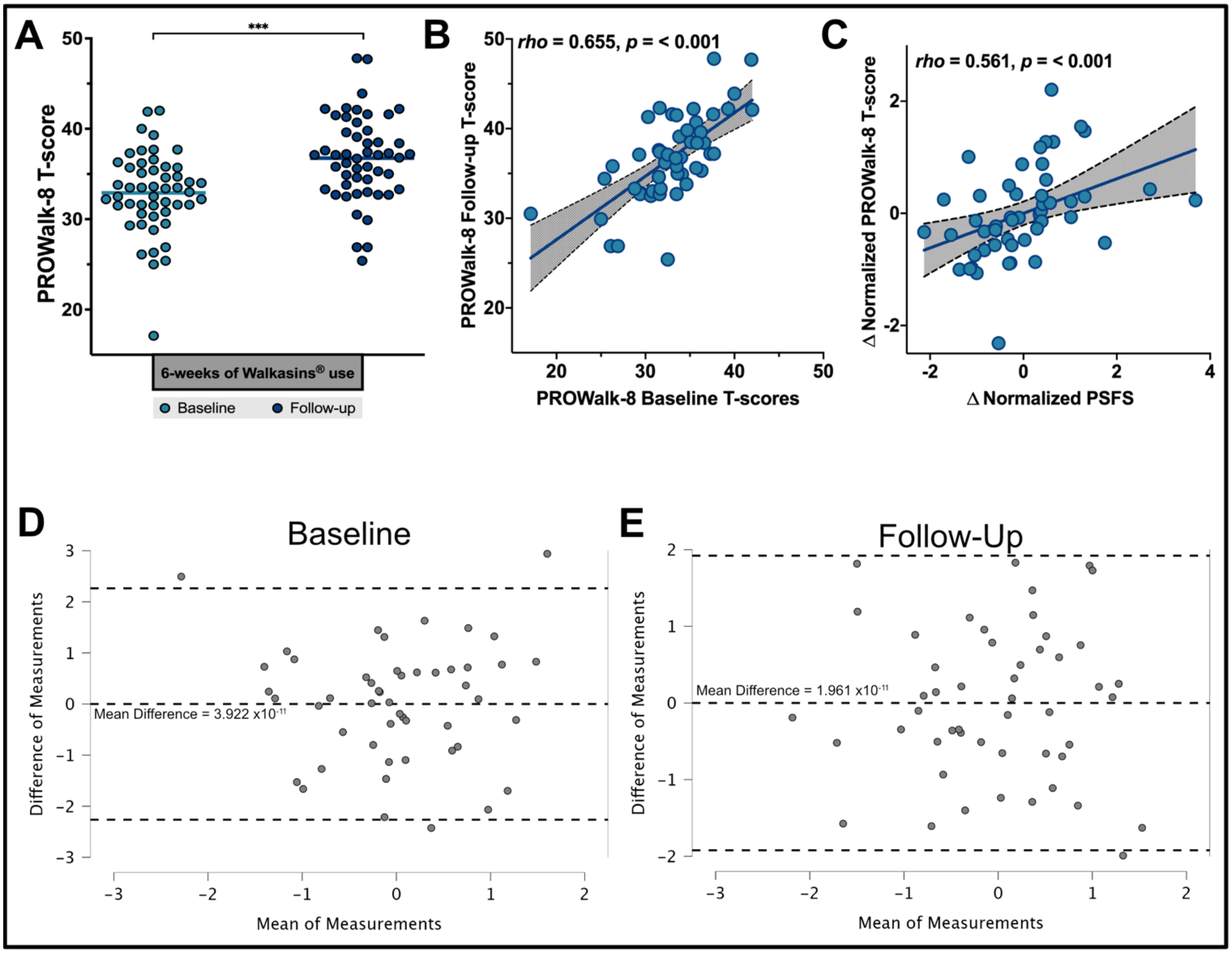


**Supplemental Figure 2. A)** PROWalk-8 performance before (32.91 ± 4.48) and six weeks after (36.75 ± 4.72) wearing Walkasins® in persons with sensory peripheral neuropathy. The triple star indicates significance at the 0.1% level. Correlations between trials and the change in scoring between instruments across the sample. Significant, positive correlations were identified for the **B)** PROWalk-8 (*rho* = 0.655, *p* = <0.001) and **C)** the change (Δ) in normalized instrument scores over time (*rho* = 0.561, *p* = <0.001). Agreement between the normalized PROWalk-8 and PSFS instruments was assessed using Bland–Altman plots at both baseline (**D**) and follow-up (**E**) timepoints.
